# JARVIS, should this study be selected for full-text screening? Performance of a Joint AI-ReViewer Interactive Screening tool for systematic reviews

**DOI:** 10.64898/2026.04.08.26350384

**Authors:** G. H. C. Barreto, C. Burke, P Davies, M. Halicka, C. Paterson, P. Swinton, B. Saunders, J.P.T. Higgins

**Affiliations:** Population Health Sciences, Bristol Medical School, University of Bristol, Bristol, UK; NIHR Bristol Evidence Synthesis Group, University of Bristol, Bristol, UK; Department of Psychology, University of Bath, Bath, UK; School of Health, Robert Gordon University, Aberdeen, UK; Applied Physiology and Nutrition Research Group – School of Physical Education and Sport and Faculdade de Medicina FMUSP, Universidade de São Paulo, São Paulo, Brazil; NIHR Applied Research Collaboration West (ARC West) at University Hospitals Bristol and Weston NHS Foundation Trust, Bristol, UK

## Abstract

**Background:** Systematic reviews are essential for evidence-based decision making in health sciences but require substantial time and resource for manual processes, particularly title and abstract screening. Recent advances in machine learning and large language models (LLMs) have demonstrated promise in accelerating screening with high recall but are often limited by modest gains in efficiency, mostly due to the absence of a generalisable stopping criterion. Here, we introduce and report preliminary findings on the performance of a novel semi-automated active learning system, JARVIS, that integrates LLM-based reasoning using the PICOS framework, neural networks-based classification, and human decision-making to facilitate abstract screening.

**Methods:** Datasets containing author-made inclusion and exclusion decisions from six published systematic reviews were used to pilot the semi-automated screening system. Model performance was evaluated across recall, specificity and area under the curve precision-recall (AUC-PR), using full-text inclusion as the ground truth. Estimated workload and financial savings were calculated by comparing total screening time and reviewer costs across manual and semi-automated scenarios.

**Results:** Across the six review datasets, recall ranged between 98.2% and 100%, and specificity ranged between 97.9% and 99.2% at the defined stopping point. Across iterations, AUC-PR values ranged between 83.8% and 100%. Compared with human-only screening, JARVIS delivered workload savings between 71.0% and 93.6%. When a single reviewer read the excluded records, workload savings ranged between 35.6 % and 46.8%.

**Conclusion:** The proposed semi-automated system substantially reduced reviewer workload while maintaining high recall, improving on previously reported approaches. Further validation in larger and more varied reviews, as well as prospective testing, is warranted.

## Background

Systematic reviews are an essential source of scientific evidence (Murad et al., 2016), and are frequently used in health sciences to inform evidence-based guidelines, decision making, and public policy (South and Lorenc, 2020). Producing a systematic review involves several time-consuming stages, many of which are performed in duplicate to support methodological rigor (Higgins and Green, 2024, Page et al., 2021) and require subject-specific knowledge (i.e., highly specialised labour) (Borah et al., 2017). As such, reviews are resource-intensive with an estimated expenditure of ∼US$140,000 per systematic review (Michelson and Reuter, 2019). One of the most time-consuming processes is screening the titles and abstracts (TA) of the records retrieved by the search against specified eligibility criteria (Haddaway and Westgate, 2019). In the context of reviews of intervention studies, the eligibility criteria are often based on the PICOS framework, which summarizes evidence in relation to ‘population’, ‘intervention’, ‘comparator’, ‘outcome’ and ‘study design’. At the TA stage, reviewers will sift potentially relevant titles and abstracts (‘Retrieve’ articles), removing obviously irrelevant records (‘Excludes’) from being fully inspected at the full-text (FT) stage of screening. At FT stage, reviewers decide on whether records are eligible for inclusion in the review (‘Includes’), removing records that do not meet these eligibility criteria.

Several key stages in the systematic review workflow have been identified as use cases for the application of automation using artificial intelligence (AI) and advanced computing technologies such as machine learning (ML) (Toth et al., 2024). Search strategies of electronic databases should maximise sensitivity, whilst aiming for reasonable precision (Higgins and Green, 2024), which often results in a high proportion of irrelevant records at the TA stage. Automation approaches can reduce the amount of time a reviewer spends reading irrelevant abstracts. ML techniques, in this context, would act as an automated classifier that learns patterns in text based on human decisions (Shemilt et al., 2014). ML-based, active learning (AL) techniques have shown promise for this task; during screening, an active learning model will progressively learn from the reviewer’s decisions, increasing their predictive performance the more information it is fed (Ferdinands et al., 2023).

The predictive performance of these methods can be measured, being frequently represented by specificity (percentage of ‘Exclude’ records correctly excluded), recall (percentage of ‘Include’ records correctly tagged as ‘Retrieve’), and area under the precision-recall curve (AUC-PR; the trade-off between accurately tagging ‘Include’ records as ‘Retrieve’ [recall] and over-including them [precision: the ratio of true positives to all predicted positives], i.e., saving time without missing records). A measure that is more specific to systematic reviews is work saved over sampling (WSS), which reflects how much workload would be saved when the active learning algorithm reaches a specific recall, usually around 95%, accepting that there would be no guarantee of a tool achieving 100% at all times (Ofori-Boateng et al., 2024). Existing ML tools, however, are mostly included in systematic screening as prioritisation tools, and therefore still require reviewers to sift through the entire set of records in a review. To date, there is no agreed criterion for a ‘safe’ threshold to stop the screening process while achieving acceptable recall (Callaghan et al., 2024).

Other rising technologies have recently been tested with the same objective, such as the use of large language models (LLMs) (Lieberum et al., 2025). These increasingly implemented tools —the most common being the generative pretrained transformer (GPT) —are deep neural network models capable of generating text. Researchers can take advantage of the LLMs’ conversational abilities by, for example, using them as second reviewers (Tran et al., 2024). It is also possible to create a prioritisation system, through a question-and-answer approach based on criteria such as PICOS, allowing reviewers to read the most relevant records first (Wang et al., 2023). However, the use of these tools is not yet commonplace among reviewers, as many are not fully convinced that they are ready for integration into systematic review workflows. The mixed findings from existing validation studies remain a major concern for full implementation (Lieberum et al., 2025).

Despite ML and LLM techniques showing promising results, challenges remain. First, many reviewers remain unconvinced that LLMs are sufficiently reliable on their own, primarily due to the risk of hallucinations (i.e., generating distorted information (Sun et al., 2024), and presenting it as if it were correct). Second, although ML tools can make screening more efficient through prioritisation, it is common for reviewers to examine the entire set of records, for no universal stopping criterion exists (Repke et al., 2026). Without such a criterion, reviewers either risk missing relevant records or experience substantially reduced time-saving benefits. A tool with a well-defined stopping criterion will enable reviewers to reduce the burden of screening even further. To meet these needs, we propose JARVIS, a semi-automated active learning screening system that combines LLM-based PICOS assessments and ML models trained exclusively on human decisions. In JARVIS, humans are the final decision-makers, and are assisted by the system’s recommended stopping criterion, further reducing screening time. This article describes JARVIS system, and reports findings from preliminary tests of performance and estimated workload savings.

## Methods

In the following sections, we outline the architecture and workflow of JARVIS, including its integration of LLM-based PICOS reasoning, priority sampling, and neural-network classification within an active-learning framework. We then describe how the system was piloted across six previously completed systematic reviews, using authors’ original screening decisions to simulate human–AI interaction during title and abstract screening. Model performance was evaluated iteratively using recall, specificity, and AUC-PR, alongside cumulative recall to capture joint human–AI accuracy (see *Performance metrics* below). We also estimated the potential reduction in reviewer workload by comparing the number of abstracts screened under manual versus semi-automated workflows.

### Overview of the JARVIS system

JARVIS is a semi-automated, active learning system that uses both generative LLMs and text-based ML for the auto-classification of abstracts according to their relevance to systematic reviews. The system’s active learning component means that it learns from human decisions as they are made, allowing it to be easily implemented in a review workflow. A summary of JARVIS’ R pipeline is shown in Figure 1.

**Figure 1.**
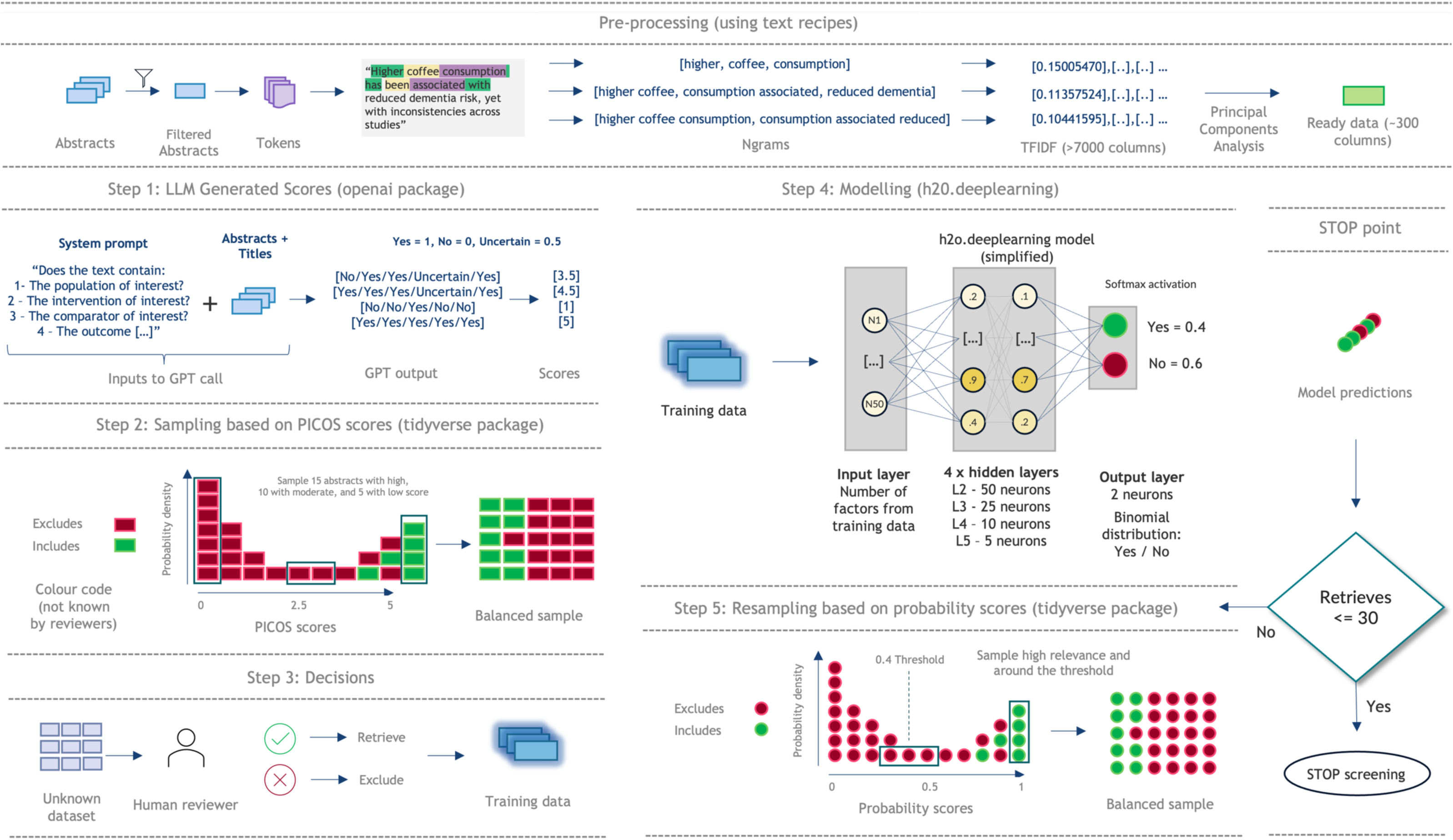
A graphical summary describing JARVIS’ step-by-step process.

#### Software

A preliminary version of JARVIS was developed and tested in R (version 4.5.2, 31-10-2025) using RStudio (2025.09.2 Build 418, Posit Software, PBC). Because the H2O package (for ML training and inferencing) requires Java, version 17 was installed (versions 8-17 are compatible). JARVIS was developed using a Dell Pro 13 Plus (Windows 11) but was fully tested in MacOS (Sequoia Version 15.7.2), which is where the results shown below were obtained. Slight differences in results (<1%) may occur due to different CPU and GPU architectures (e.g., Apple Silicon, Intel or Nvidia GPUs).

#### Text data pre-processing

Initially, only the abstracts are converted into variables the ML model can use as predictors. Our preliminary tests indicated that including titles provided negligible additional information for the ML models and they were therefore omitted. The entire corpus of abstracts belonging to the review is tokenised and filtered for stop words based on the SMART list (System for the Mechanical Analysis and Retrieval of Text) (Rocchio, 1971). Stop words are those considered to have little semantic relevance, such as articles, prepositions, conjunctions, and pronouns. The remaining tokens are then combined into ‘n-grams’ (sequences of 1 to 3 words), with a set limit of combinations (7000 most relevant terms). The Term Frequency-Inverse Document Frequency (TF-IDF) (Salton and Buckley, 1988) is then calculated for each of the ngrams, converting the text data into a continuous numerical format (their relative frequency in each abstract) suitable for modelling. To reduce the dataset dimensionality, the TF-IDFs (7,000 columns) are condensed through a Principal Components Analysis (PCA). We retained 90% of the variance of the TF-IDF columns. All the text processing was done with the *textrecipes* package (Hvitfeldt et al., 2025).

### Step 1: LLM-generated scores

The method uses the *create_chat_completion* function (temperature = 0.1, max_tokens = 300) of the *openai* package (Rudnytskyi, 2023) to obtain responses from any available OpenAI model via API (Application Programming Interface). All instructions are sent as a system prompt, including the “role” the bot must play, the reported PICOS elements and PICOS-related questions (e.g., “1 - does this study report on the population of interest? 2 - does this study report on the intervention of interest? […]”). An additional question is included – “Does the abstract below represent a literature review?” – aiming to improve the categorisation even further. The responses to the questions are limited (via prompt) to “Yes”, “No”, or “Uncertain”. For each interaction with the API, one title and one abstract, unmodified, are sent as the main user prompt, to which the model responds in the desired format. The instructions also suggest a separator to facilitate breaking the responses into columns (e.g., “No/Yes/Yes/No/No/Yes”). For the next step (sampling with priority), the responses are converted into a score. Except for the “review” question, each “Yes” response receives 1 point, and every “Uncertain” and “No” responses receive 0.5 and 0 points, respectively. The possible scores, therefore, range between 0 and 5.

### Step 2: Initial sampling (PICOS-based)

Instead of sampling at random, this AI-assisted system allows for priority sampling from the beginning based on the PICOS score. For this initial step, 15 top scoring records (score 5) are selected, along with 10 mid-range (score 2.5) and 5 low scoring records (score 0). This allows for a greater number of potentially relevant records to be selected for the first iteration, improving sample balancing.

### Step 3: Decisions

At this stage, the 30 sampled records are given to human reviewers for decision-making (‘Retrieve’/‘Exclude’). For this, reviewers should use information available in titles and abstracts. The abstracts with human decisions are then used in Step 4 to train the machine learning models.

### Step 4: Modelling

The ML models (feed-forward artificial neural networks; *h2o.deeplearning* from the *h2o* package) (Fryda et al., 2024) are trained on-the-go based on the decisions made by human reviewers (y variable), while the predictors (x variables) are the text extracted variables (condensed TF-IDFs) and the dummy variable responses to the PICOS questions (Yes: 0, 1; No: 0, 1; Uncertain: 0, 1). While it is usually advisable to split data into training/validation/test sets, there are not as many data points when a new review is initiated. Here, a modified version of the EasyEnsemble method was used (Liu, 2009). This method creates three separate models (ensembles), each trained on a full replica of the minority class (‘Retrieves’) paired with a random sample of the majority class (‘Excludes’). These training samples are obtained from the records with human decisions at the specific point (n = 30 * round number, see Step 5: Iterative sampling). For JARVIS, the three new samples are manually tagged as folds (k-folding), and h2o is instructed to use them to cross-validate each other. Instead of using the core generated model, all three k-models are used to create predictions of probability for the remaining abstracts, representing how likely they are to be selected. Each unread abstract has, then, three separate predictions that are averaged to define the final probability. Those with final probability above 0.4 are classified as ‘Retrieves’, while the remaining are ‘Excludes’. This threshold (0.4 instead of the standard 0.5) was chosen specifically to be more permissive in classification, aiming to obtain higher recall.

Hyperparameter optimisation was done in two steps. Initially, exploratory trials were run, aiming to identify a stable region. An iterative grid search was then conducted, evaluating each parameter combination across all review datasets and across the full sequence of iterations. This procedure allowed us to identify configurations that generalised well across topics and maintained high recall throughout the active-learning process. All training-sampling steps used a fixed random seed (123) to minimise stochastic variation and ensure reproducibility. The hyperparameters of the h2o model are presented in Table 1.

**Table 1.** The final hyperparameter configuration for the h2o deep learning models used.

| Hyperparameter | Value |
| --- | --- |
| Cross-validation | k-folds: 3 user-created folds |
| Initial weight distribution | Uniform Adaptive |
| Gradient Descent | Nesterov |
| Seed | 123 |
| Layers | 4 |
| Neurons per layer | 50, 25, 10, 5 |
| Activation at hidden layers | Tanh |
| Activation at output layer | Automatic (SoftMax) |
| Max Epochs | 100 |
| Stop rounds | 3 |
| Stop tolerance | 1e-5 |
| Stop metric | Mean per class error |
| Learning rate | 0.001 |
| Rate annealing | 0.001 |
| Loss function | Cross Entropy |
| User-controlled weights | 40 for 'Retrieves', 1 for 'Excludes' |
| L1 regularisation | 0 |
| L2 regularisation | 0.02575 |

### Step 5: Iterative sampling (model-based)

Since it is unlikely that all studies are classified correctly after only 30 human decisions, we suggest that Steps 2 - 4 are repeated iteratively. For this resampling stage, instead of based on the PICOS score, the sampling is done based on the probabilities generated by the model. This time, 15 of the records with the highest probability are selected, as well as 15 sitting around the threshold value (8 below and 7 above), and then given to human reviewers for decision-making. At each iteration, the 30 new abstracts with decisions join the previous ones and a new model is trained. This way, the models are trained with a more varied sample which would lead to better generalisation. This process is to be repeated until no more than 30 records (or 1% of the total of abstracts, whichever was lower) remain above the threshold (STOP point). Ideally, as our tests have shown, the user will have reached approximately 100% recall at this point, minimising the loss of relevant studies.

### Evaluation using archived systematic review datasets

Three published systematic reviews and meta-analyses (SRMAs) (Barreto et al., 2023, Jackson et al., 2019, Pickard et al., 2014), and two network-meta-analyses (NMA) (Sterne et al., 2017, Webster et al., 2025) were selected for pilot testing. Sterne et al. (2017) included two research questions with separate search strategies and PICOS elements and was therefore considered as two separate reviews for the purposes of testing. The PICOS and aims of each of the included reviews are described in Table 2. The reviews were selected based on accessibility for pilot data testing and cover a wide range of health science-related topics. All selected reviews were performed with the highest level of rigour, including dual double-blinded TA and FT screening. Necessary information was obtained from one of the authors of each review as a spreadsheet in .xlsx or .csv format. The files contained, at minimum: (i) the titles and abstracts of all the search results, (ii) reviewer-made title/abstract decisions (Retrieves/Excludes), and (iii) reviewer-made full-text decisions (Includes/Excludes). Records with no abstracts were excluded from the datasets. The total number of records in each review, as well as the number of records without an abstract, ‘Retrieves’, ‘Excludes’ at TA, ‘Includes’ and ‘Excludes at FT’ are described in Table 3. The reasons for exclusion by human reviewers were not detailed at title/abstract stage, as is standard practice.

**Table 2.**
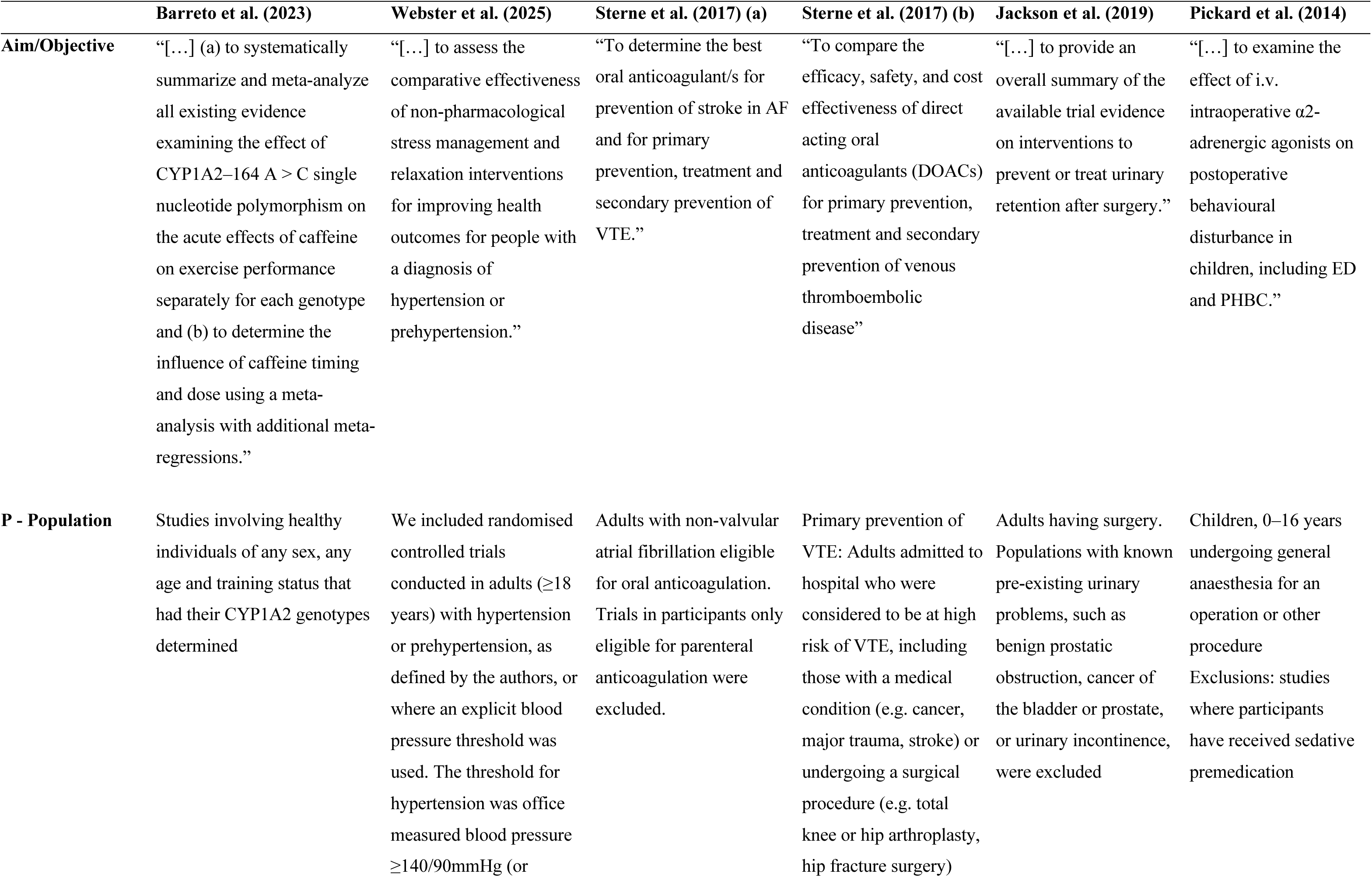
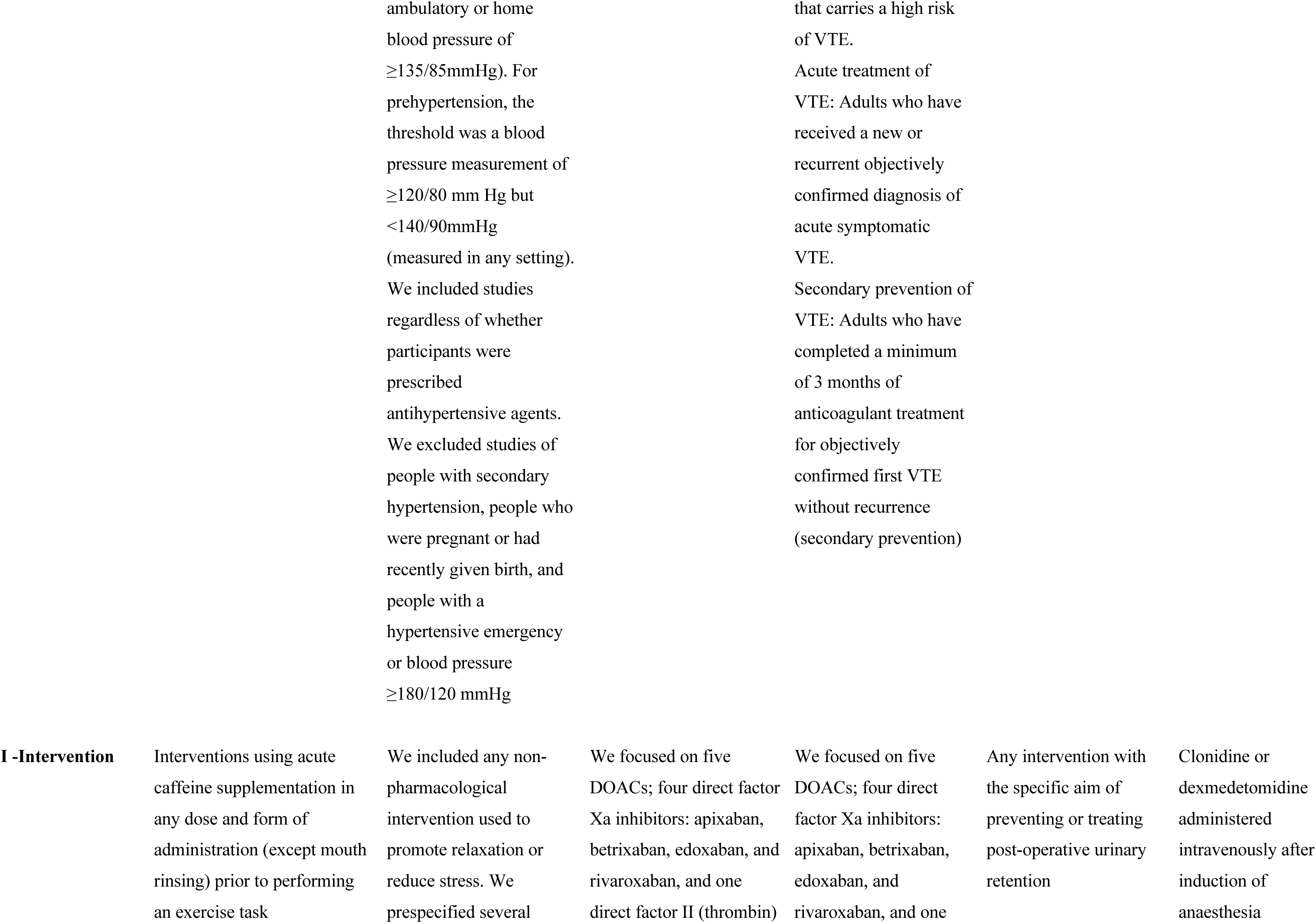
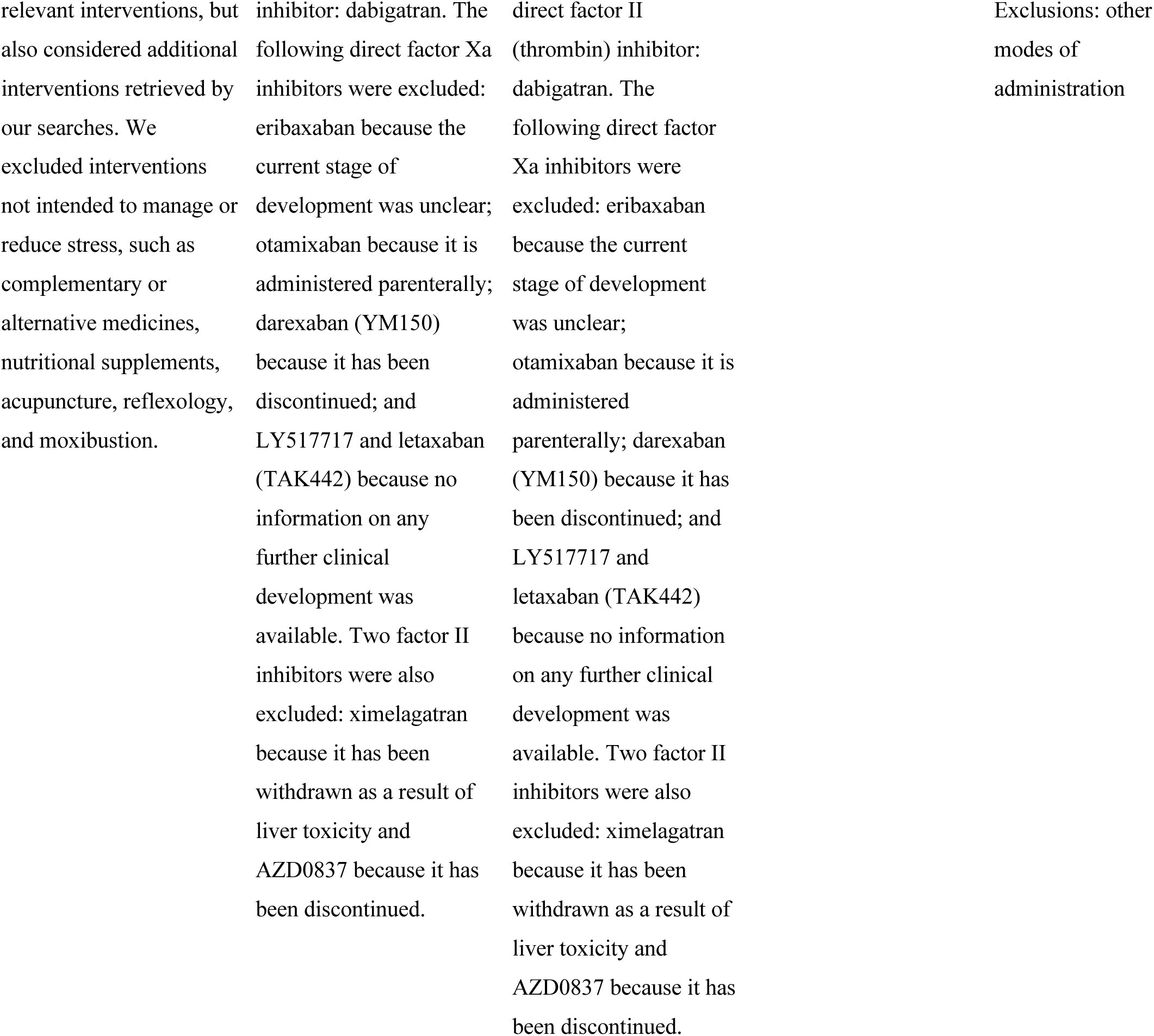
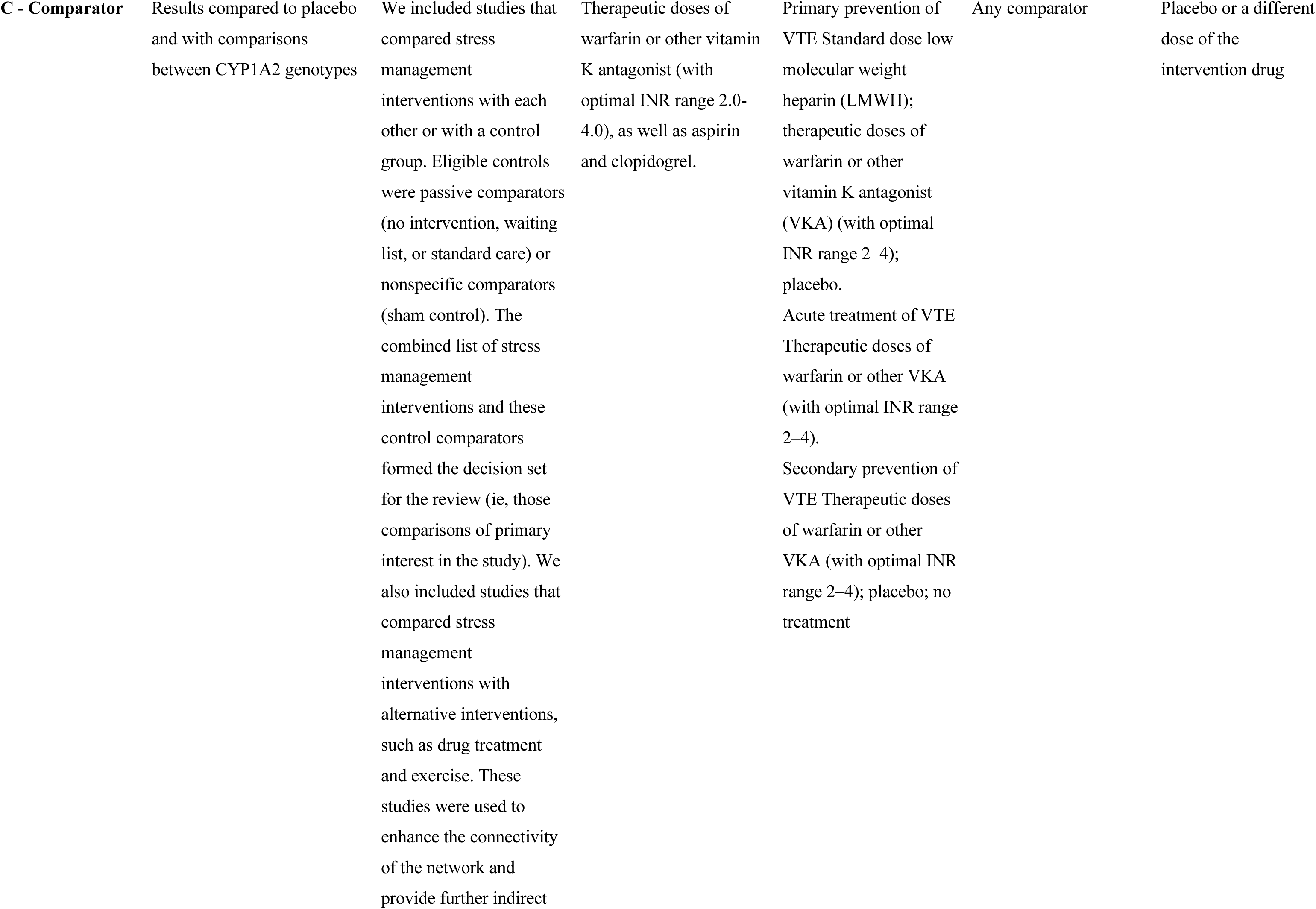
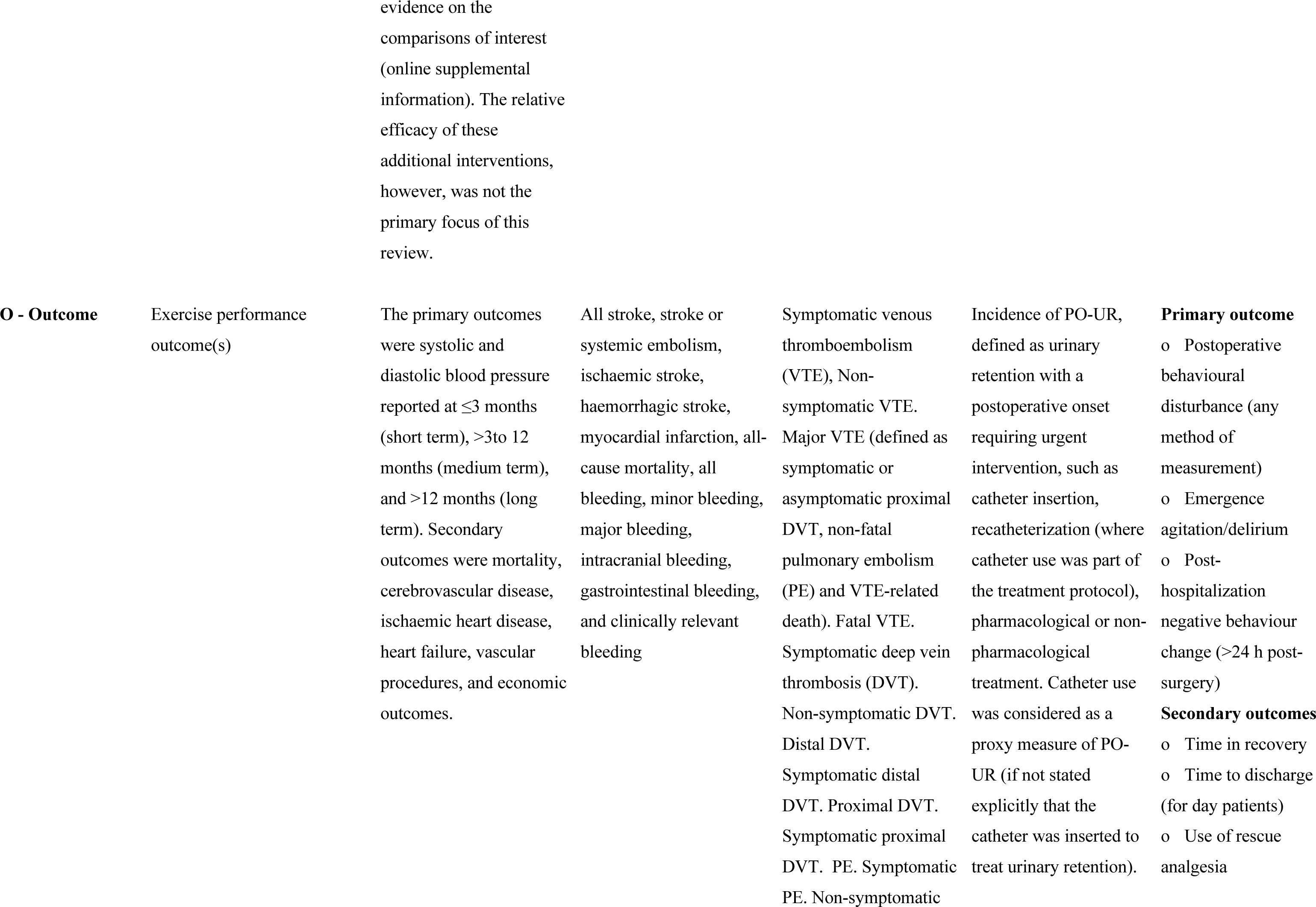
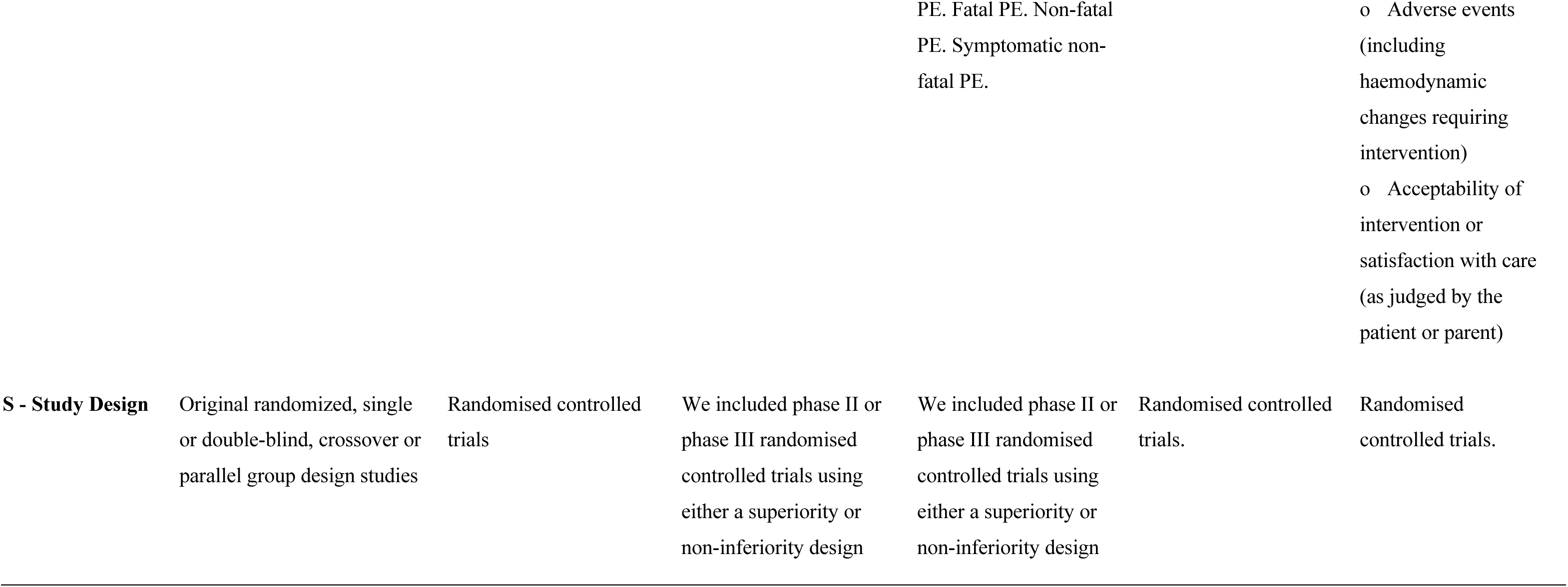
PICOS for each archived systematic review dataset included in the evaluation (as defined by the original authors).

**Table 3.** Number of records at each stage of screening for the included reviews. Retrieves are records that went to FT screening, while Includes are studies that made it to the final review.

|  | Total<br>records | Records with<br>abstracts | n Retrieves<br>(%) | n Includes (%) |
| --- | --- | --- | --- | --- |
| *Barreto et al. (2023)* | 807 | 807 | 20 (2.48) | 19 (2.35) |
| *Webster et al. (2025)* | 7452 | 7451 | 429 (5.76) | 193 (2.59) |
| *Sterne et al. (2017) (a)* | 1852 | 1632 | 161 (9.87) | 102 (6.25) |
| *Sterne et al. (2017) (b)* | 2726 | 2420 | 194 (8.02) | 110 (4.55) |
| *Jackson et al. (2019)* | 2140 | 2050 | 181 (8.82) | 22 (1.07) |
| *Pickard et al. (2014)* | 607 | 574 | 20 (3.48) | 12 (2.09) |

The OpenAI model used for these experiments was the “gpt-4.1-mini”, and its responses to the PICOS and review questions were obtained as described above. The initial sampling was performed based on the PICOS score and the decision process was simulated by extracting the original authors’ double-screened title/abstract decisions. These decisions were then used as y variables to train the model, and new predictions were created until the **STOP point** was reached. R codes for tests and validations of this system are available online (https://github.com/gabsbarreto/JARVIS-R).

#### Performance metrics

To assess the predictive performance of the models, values of recall, specificity, and AUC-PR were calculated for each iteration (Iteration recall, Specificity, AUC-PR). In this case, these were calculated based on the proportion of final FT decisions of ‘Include’ and ‘Exclude’ at each iteration on unseen records (i.e., those that have not yet been sampled). Joint recall (or cumulative recall) was calculated based on the combination of positive cases identified by the reviewer (seen) and the positive cases correctly classified by JARVIS. This measure aims to evaluate the performance of the human-AI integrated workflow. The choice to use FT decisions aimed to measure loss and recall of records that were confirmed to be 100% relevant to the review. AUC-PR was calculated based on the integral of precision in function of recall with the *pr.curve* function of the *PRROC* package (Grau et al., 2015). Since AUC-PR was not manually calculated, the formulae are not shown here. Recall, cumulative recall and specificity are given by:

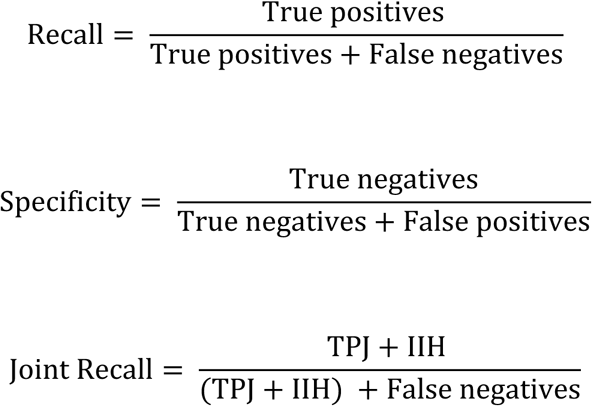

where TPJ represents the true positive cases according to the model’s classification, and IIH represents the total of FT ‘Includes’ identified by the human reviewers.

Workload savings were defined as the percentage of abstracts reviewers would not have to read in the case JARVIS was used, at the threshold of 95% of ‘Includes’ identified by the human reviewer. However, since JARVIS employs batch sampling, the first iteration at which the threshold was reached was used to calculate WSS@95%. Workload savings at JARVIS’ STOP point were also calculated (WSS@STOP). Effective WSS@STOP was also calculated, consisting of the total time saved if a single reviewer still screens the abstracts JARVIS classified as ‘Excludes’.

## Results

For each review, the density distributions of predicted probabilities are presented (Figures 2-7). The progression of the % of ‘Includes’ captured, iteration recall, joint AI-reviewer recall, and specificity are presented in Figure 8. Workload savings are presented in Table 4.

**Figure 2.**
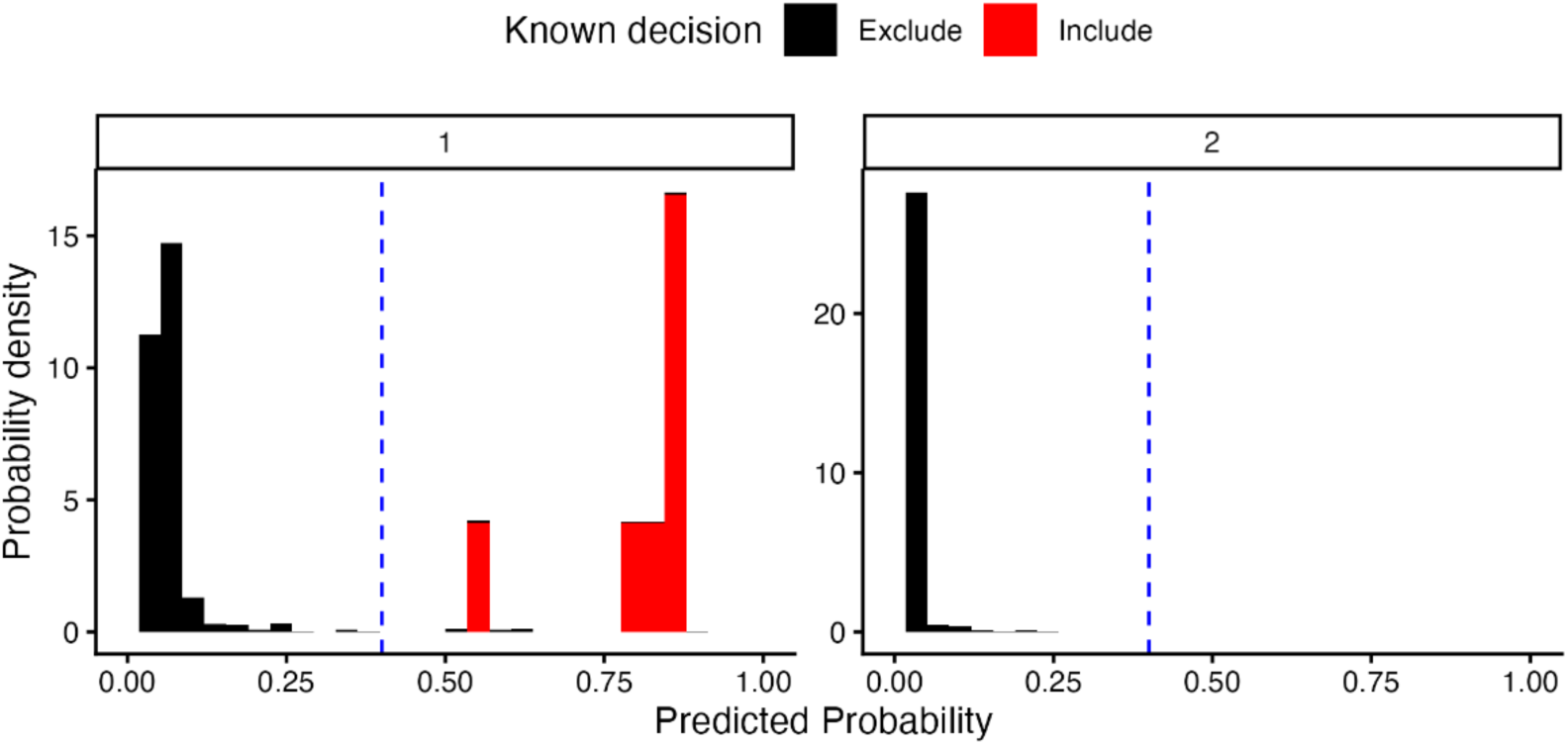
Density distribution of predicted probabilities for known ‘Includes’and ‘Excludes’ at the first (1) and final (2) iterations for Review 1. The classification threshold of 0.4 is indicated by the blue dashed lines.

**Figure 3.**
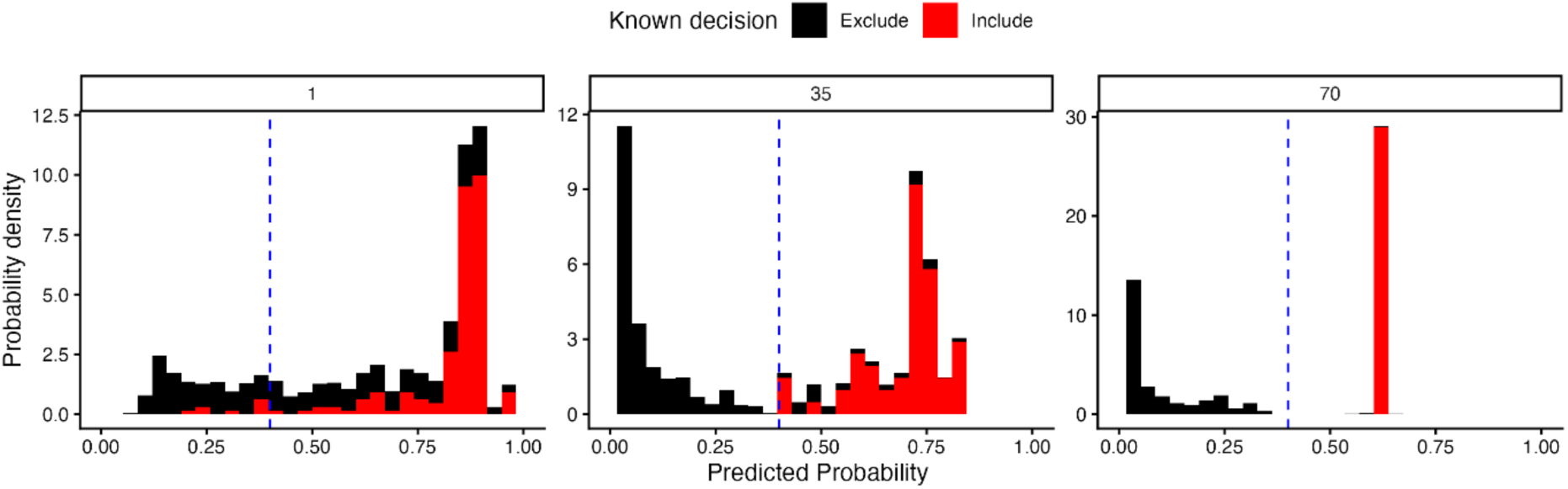
The density distribution of predicted probabilities separated between known ‘Includes’ and ‘Excludes’ at the first (1), middle (35), and final (70) iterations for Review 3. The classification threshold of 0.4 is represented by the blue dashed lines.

**Figure 4.**
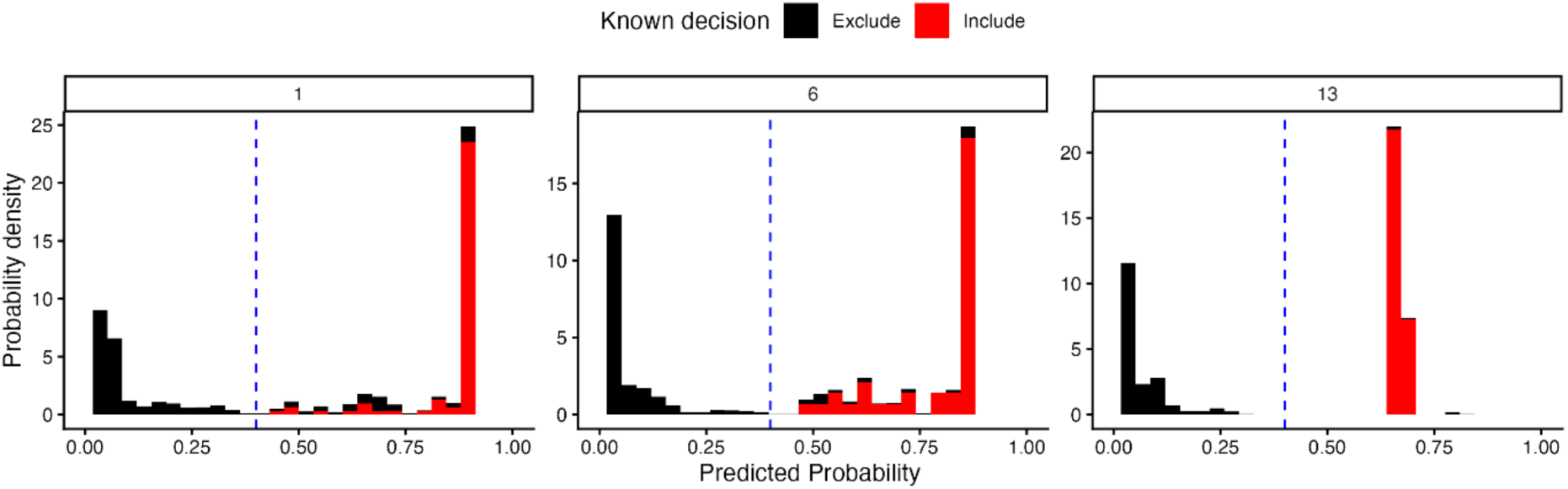
The density distribution of predicted probabilities separated between known ‘Includes’ and ‘Excludes’ at the first (1), middle (6), and final (13) iterations for Review 3. The classification threshold of 0.4 is represented by the blue dashed lines.

**Figure 5.**
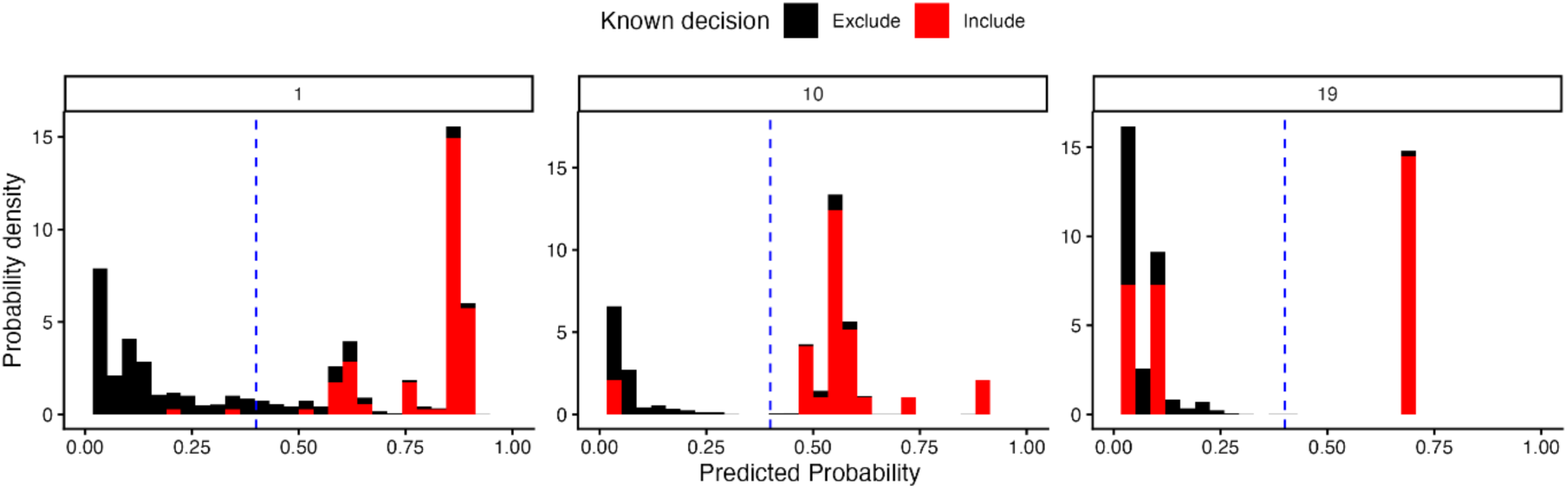
The density distribution of predicted probabilities separated between known ‘Includes’ and ‘Excludes’ at the first (1), middle (10), and final (19) iterations for Review 4. The classification threshold of 0.4 is represented by the blue dashed lines.

**Figure 6.**
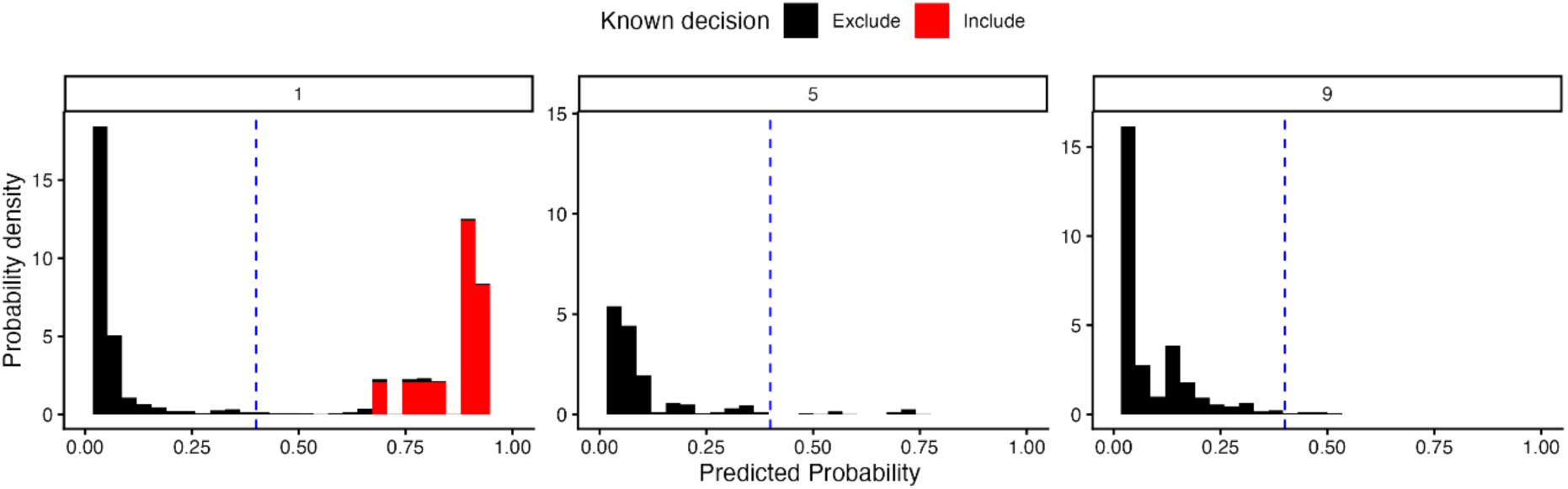
The density distribution of predicted probabilities separated between known ‘Includes’ and ‘Excludes’ at the first (1), middle (5), and final (9) iterations for Review 5. The classification threshold of 0.4 is represented by the blue dashed lines.

**Figure 7.**
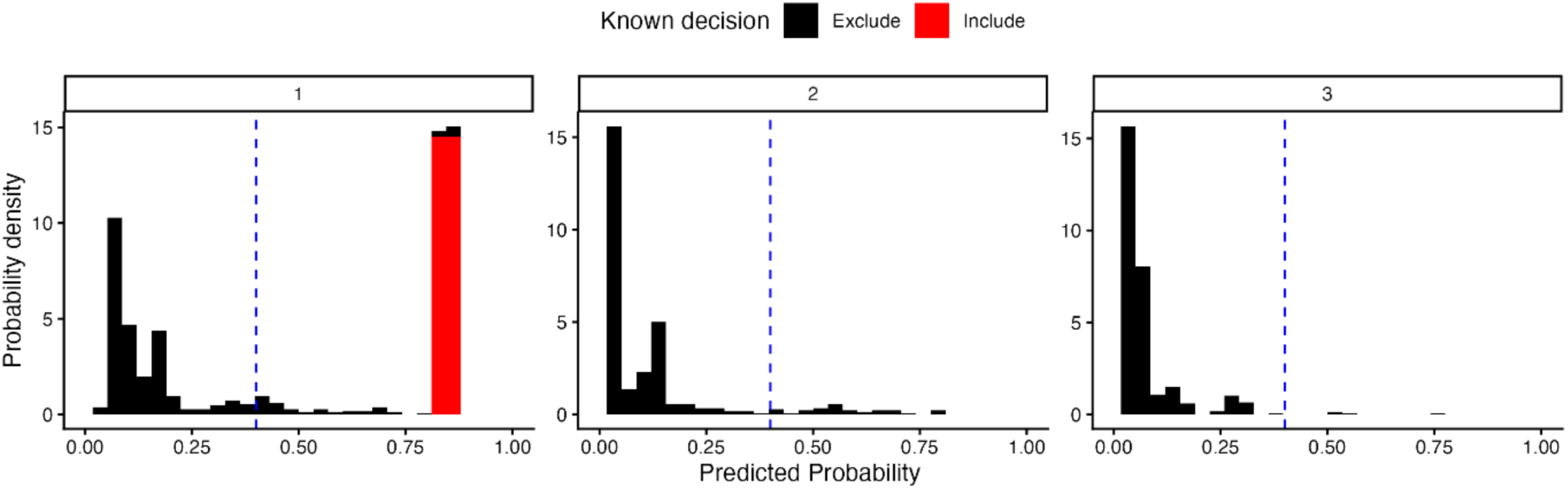
The density distribution of predicted probabilities separated between known ‘Includes’ and ‘Excludes’ at the first (1), middle (2), and final (3) iterations for Review 6. The classification threshold of 0.4 is represented by the blue dashed lines.

**Figure 8.**
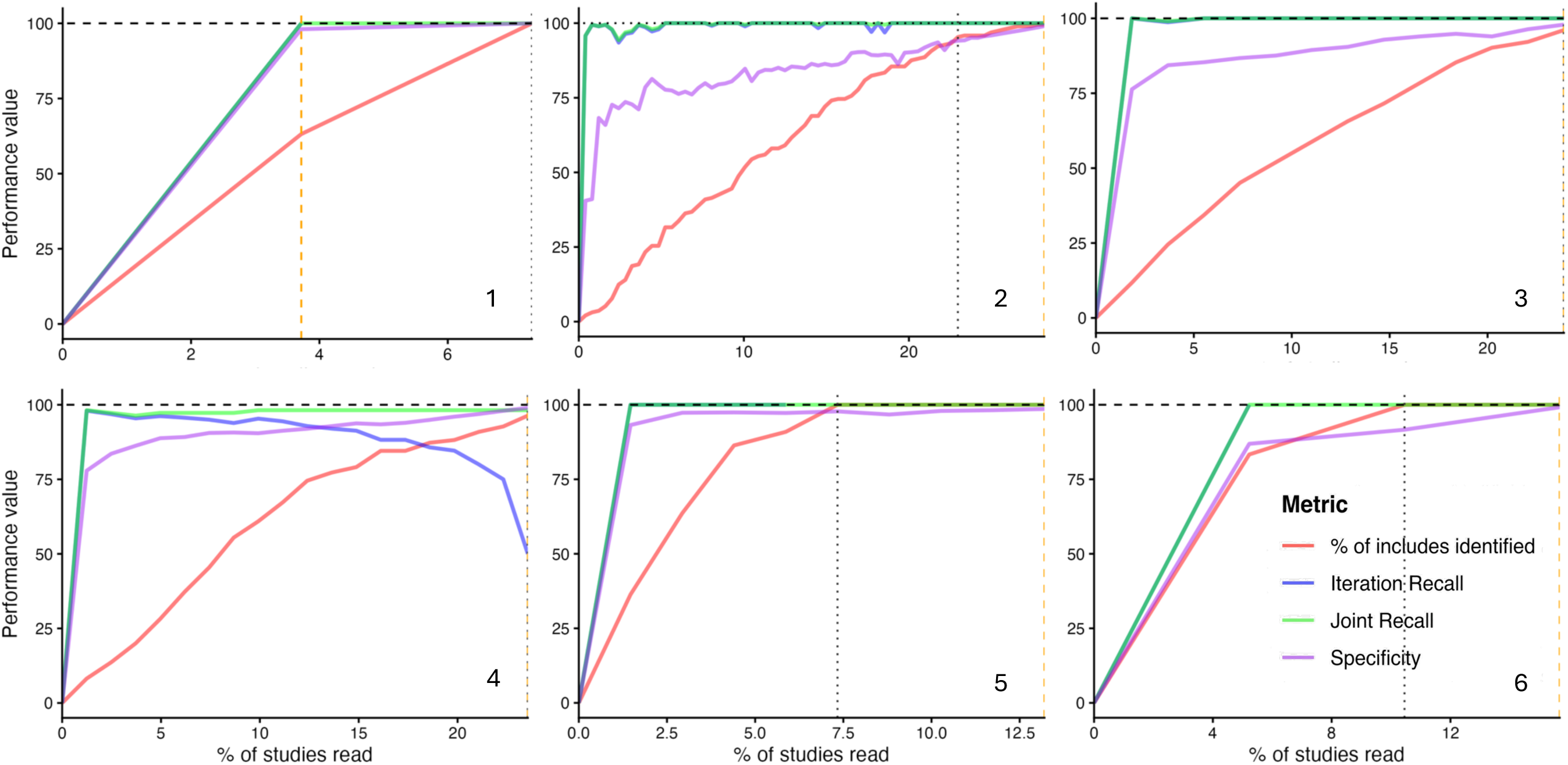
The progression of the percentage of ‘Includes’ identified, within-iteration recall, joint JARVIS-reviewer recall, and specificity as a function of the proportion of records screened by human reviewers for reviews 1) Barreto et al. (2023), 2) Webster et al. (2025), 3) Sterne et al. (2017) (a), 4) Sterne et al. (2017) (b), 5) Jackson et al. (2019) and 6) Pickard et al. (2014). The orange dashed line represents the STOP point, while the dotted line represents the moment at which 95% of ‘Includes’ were identified. These lines overlap if WSS@95% was achieved on the same iteration. Where iteration recall and joint recall are identical, they are superimposed.

**Table 4.** Total workload savings at the TA screening stage estimated using JARVIS across the six archived systematic review datasets.

| Indicator | Review 1<br>(Barreto et al.,<br>2023) | Review 2<br>(Webster et al.,<br>2025) | Review 3<br>(Sterne et al.,<br>2017) (a) | Review 4<br>(Sterne et al.,<br>2017) (b) | Review 5<br>(Jackson et al.,<br>2019) | Review 6<br>(Pickard et al.,<br>2014) |
| --- | --- | --- | --- | --- | --- | --- |
| Total records | 807 | 7,446 | 1,632 | 2,420 | 2,045 | 574 |
| Records read at<br>STOP point | 30 | 2,100 | 361 | 570 | 270 | 90 |
| WSS@95% | 92.7% | 72.4% | 74.3% | 75.5% | 90.6% | 82.1% |
| WSS@STOP% | 93.6% | 71.1% | 74.3% | 75.5% | 85.5% | 83.6% |
| Effective<br>WSS@STOP% | 46.8% | 35.6% | 37.2% | 37.8% | 42.8% | 41.8% |

STOP point, the moment in which JARVIS suggests there are less than 30 records (or 1% of the total, whichever is lower) to screen; WSS@95%, work saved over sampling at 95% recall; WSS@STOP% work saved over sampling at JARVIS STOP point; Effective WSS@STOP%, all records would be screened, though only one human reviewer would read those excluded by JARVIS.

### Review 1 – Barreto et al. (2023)

Only one round was necessary to reach the STOP point for review 1 (Figure 8, Panel 1, orange dashed line), representing 3.7% of the 807 records. We extended the analysis to a second round to reach the WSS@95% threshold. At the STOP point, 63.2% of the ‘Includes’ had been identified by human reviewers, while the remaining were correctly classified by JARVIS (joint and iteration recall 100%) at a specificity of 98.1%. Workload savings at the STOP point, therefore, were 93.6%, decreasing to 92.7% after the identification of at least 95% of the ‘Includes’ (WSS@95%, Figure 8, dotted line). AUC-PR values ranged from 95.9 to 100%, with a mean of 97.8%.

### Review 2 – Webster et al. (2025)

For the second review, 70 rounds (28.2% of 7446 records) were necessary to reach the STOP point (Figure 8, Panel 2, orange dashed line), with 99.5% of ‘Includes’ identified. The remaining record was correctly classified by JARVIS (Figure 3, iteration 70), with both within and joint JARVIS-reviewer recall of 100% at a specificity of 99.0%. A WSS@95% of 72.4% was found, reducing to 71.1% after reaching the STOP point. AUC-PR values ranged from 92.1 to 99.9%, with a mean of 96.5%.

### Review 3 – Sterne et al. (2017) (a)

For the third review, 13 rounds were necessary to reach the STOP point, representing 22.1% reads out of the 1632 records (Figure 8, Panel 3, orange dashed line). At that point, both within iteration and joint recalls were at 100%, with 97.9% specificity. The 95% threshold for the WSS@95% was reached at the STOP point (96.1% of ‘Includes’ identified), with 74.3% estimated workload savings at that point. AUC-PR values ranged from 83.8 to 98.7%, with a mean of 91.6%.

### Review 4 – Sterne et al. (2017) (b)

For the fourth review, 19 rounds were necessary to reach the STOP point, representing 23.6% reads out of the 2420 records (Figure 8 Panel 4, orange dashed line). At the same point, joint recall obtained was 98.2%, while iteration recall was 50%. This change occurred due to the difference in prevalence of ‘Includes’ in the remaining sample (n = 2). The 95% threshold for the WSS@95% was reached at the STOP point (96.4% of ‘Includes’ identified), with 75.5% estimated workload savings at that point. AUC-PR values ranged from 87.4 to 99.1%, with a mean of 93.97%.

### Review 5 – Jackson et al. (2019)

For the fourth review, 9 rounds (13.2% reads out of 2045 records) were necessary to reach the STOP point (Figure 8, Panel 5, orange dashed line). All the ‘Includes’ had been identified at that point, with 100% recall both within iteration and joint JARVIS-reviewer, and specificity of 98.6%. The WSS@95% obtained was of 90.6%, reducing to 85.5% at the stop point (WSS@STOP). AUC-PR values ranged from 96.9 to 100%, with a mean of 99.3%.

### Review 6 – Pickard et al. (2014)

For the fifth review, 3 rounds (15.7% reads out of 574 records) were necessary to reach the STOP point (Figure 8, Panel 6, orange dashed line). All ‘Includes’ had been identified at that point, with 100% recall both within iteration and joint JARVIS-reviewer, and specificity of 99.2%. The WSS@95% obtained was of 82.1%, increasing to 83.6% at WSS@STOP. AUC-PR values ranged from 98.6 to 100%, with a mean of 99.5%.

## Discussion

Our evaluations suggest that using JARVIS for title and abstract screening could save, on average, 81.3% (range: 72.4 to 92.7%) of working hours associated with abstract screening for systematic reviews, with near perfect recall (range: 98.2 to 100%). If reviewers choose to screen the records classified as ‘Excludes’ at the STOP point, the effective workload savings range between 35.6 and 46.8%. If future tests confirm, however, that recall rates as high as those reported here are reproducible, we would advise systematic reviewers that continuing screening after the STOP point is likely to be unnecessary. In the largest evaluation ran so far (Webster et al., 2025), we demonstrated that a total of 71.1% of workload (i.e., from 126 to 38 hours) could be saved with this workflow. Only 2100 out of the 7446 studies had to be read/decided for to reach the STOP point, with the resulting recall value of 100%. The lowest recall obtained (98.2%), however, signals that adaptations should be implemented when reviews are more complex than a simple SRMA of RCTs.

It could be hypothesized that review complexity influenced our results. The three studies that had false negatives at some point in the process were NMAs (Webster et al., 2025, Sterne et al., 2017) (a and b). In these cases, the intervention and comparator eligibility criteria were broader than for systematic reviews with pairwise meta-analyses, which only include direct comparisons between interventions of interest and predetermined comparators. NMAs combine direct and indirect evidence to estimate relative intervention effects. Therefore, in addition to first-order comparisons (interventions of interest), NMAs often include second-order comparisons (interventions that have been directly compared with interventions of interest, as in Webster et al. [2025]), or even third-order comparisons (interventions that have been directly compared to second-order comparators, as in Sterne et al. [2017]) to draw from indirect evidence (Hawkins et al., 2009). For example, for Sterne et al. (2017), a “No” response from the LLM regarding the intervention element would have reduced the chances of a study of being selected by the model. For all other cases, the PICOS elements were more narrowly defined with either a single type of intervention or a single type of outcome. Therefore, although the results are promising, improvements may be required for more complex scenarios. For NMAs, for example, prompts should mention specifically the type of review being performed and, if third-order comparators are included, adjusting the score system to represent these studies adequately.

On the other hand, the proportion of studies classified as ‘Retrieves’ during the screening phase seems to be correlated with time savings, although no analysis could be done to confirm this. As it is currently understood, human reviewers should be over-inclusive aiming to reduce loss to a minimum (O’Mara-Eves et al., 2015). However, even though the tool has shown to lead to workload savings in this retrospective analysis, it is possible that lower retrieval rates from human reviewers could yield even better results. Further trials should test whether JARVIS’ prospective workflow can also influence reviewers’ decision-making, reducing hyper-retrieval, and if this is likely to affect results.

An advantage of the proposed system is that all neural network models used the same hyperparameters and are tuned to generalise to a wide range of reviews. This allows for minimal end-user input, who would only need to provide the PICOS elements for the review to start the process, and to make the decisions after each sampling round. For that reason, reviewers will also benefit from using JARVIS when performing rapid reviews. In addition, JARVIS models can also be stored and used to make inferences on datasets other than the ones it was trained on. In living systematic reviews, for example, reviewers would be able to load STOP point JARVIS models and use them to make inferences for the new updates. Using the checkpoint system in *h2o*, it is also possible to continue training JARVIS with the new data, further increasing its performance for new updates.

### Limitations

Despite the clear benefits of using this method, limitations exist. Firstly, this method was tested solely on systematic reviews of the effects of interventions, which is what the PICOS framework was designed for. Furthermore, the method was applied only in examples with randomised controlled trials, one of the easiest study designs to identify. The performance of the approach for other study designs and other types of research questions is not known. In reviews of other types of evidence such as exposure studies (PECOS – population, exposure, comparator, outcome and study design) or prevalence (PC – population, condition), the prompts can easily be adapted to match different frameworks but will also require new validation studies testing different combinations of hyperparameters. Nonetheless, SRMAs of interventions are the most common type of SRMA (Tufanaru et al., 2015).

In parallel, it is necessary to further validate these results with more complex review designs, such as NMAs, since only three were included in this study. For example, some NMAs consider relevant studies making comparisons that are not directly addressing the review question but contribute indirect evidence to the comparisons of interest. If the interventions in these additional studies cannot be pre-specified, this may affect how the LLMs respond to the intervention and comparator question. This is not too dissimilar to SRMAs that do not pre-specify the full list of eligible interventions in advance [e.g., Webster et al. (2025)]. Except, in this case, only one of the five elements of PICOS is affected. On the other hand, since the decisions that train the models are made by human reviewers, we expect that the neural network model training process will also capture these patterns, including different configurations of PICOS. Although the results obtained in our evaluations with NMAs were promising (98.18-100% recall and a minimum of 71% workload savings), more studies should be performed to validate JARVIS on this type of review.

Thirdly, this workflow was not validated for search hits that do not have an abstract. This is a relatively common finding in database searches, and we recommend authors make these decisions separately, adding some time to their workloads. This process could also be made easier by requesting an LLM to respond to the PICOS questions based solely on the titles. A prioritisation system could be put in place for title-only records, i.e., ranking the abstracts in a descending order according to their relevance, potentially reducing workload. However, this is beyond the scope of this paper.

Fourthly, we adopted the PICOS framework as it provides a convenient and general set of considerations commonly used to inform eligibility criteria for systematic reviews of interventions. In practice, eligibility criteria may not cover all PICOS elements (specification of outcomes is often omitted) and eligibility criteria often include additional considerations around reporting characteristics. This reflects a limitation of the common misrepresentation of PICOS as *eligibility criteria* rather than a framework to structure review *research questions*. To comprehensively cover common elements of eligibility criteria applied in systematic reviews such as language, publication type, year of publication, the PICOS framework could be extended to capture these reporting characteristics.

Finally, it is important to reiterate that this is a retrospective evaluation study carried out with datasets of abstracts that have already been categorised by humans. The precision and functionality of JARVIS still need to be assessed in prospective settings, allowing for better comparisons with existing tools. For example, several widely available abstract screening tools using text-mining ML classifiers have been previously assessed. Rayyan, for example, is estimated to help save, on average, 49% of total workload at a recall of 95% (Ouzzani et al., 2016). When tested in a real-life scenario with human reviewers, it has been shown to lead to workload savings of ∼25% at similar recall (range: 92-100%) (Li et al., 2023). Other tools have also achieved higher workload savings in the same test, averaging 73% (Abstrackr) and 77% (SWIFT-review) with recall identical to Rayyan’s (Li et al., 2023). In our evaluations, JARVIS enabled an average of 81.3% in workload savings with a near perfect recall (98.18 - 100%). The high recall achieved makes JARVIS’ workflow more than a prioritisation method, as it is the case for the other tools. It has the potential to allow reviewers to use JARVIS’ predictions as a definitive classification, further reducing workload.

## Conclusions

JARVIS can enable systematic reviewers to save, on average, 81% of workload during title and abstract screening while achieving recall values approaching 100%. These performance metrics exceed those typically reported for existing machine-learning and LLM-assisted screening tools. Based on these preliminary findings, JARVIS appears to be a feasible and safe system for TA screening. However, it should be tested in more demanding scenarios—such as reviews with >10,000 search hits and more complex eligibility structures—and evaluated in prospective workflows before its STOP criterion is used as a formal decision point.

## Data Availability

All data and validation code are available online, as described in the manuscript.

https://github.com/gabsbarreto/JARVIS-R

## Acknowledgments

Our thanks to review authors for sharing their data with the team for tests and validation. Additionally, thanks to George Spooner for assistance in articulating the model process and generating the schematics in Figure 1.

## Credit statement

Conceptualization: GB; Data Curation: GB, PD; Formal analysis: GB; Investigation: GB; Methodology: GB; Project administration: GB; Software: GB; Supervision: BS, JPTH; Validation: GB; Visualization: GB; Writing – Original draft: GB, CB, MH, CP; Writing – reviewing and editing: GB, CB, PD, MH, CP, PS, BS, JPTH

